# Mathematical models for influenza vaccination in homeless hostels

**DOI:** 10.64898/2026.07.10.26357528

**Authors:** Jingsi Xu, Natalie Hutchinson, Thomas House, Lorenzo Pellis, Andrew Hayward, Ian Hall

**Author notes:** Ian Hall, School of Mathematics, The University of Manchester, Oxford Road, Manchester M13 9PL, United Kingdom. co-first authors (Jingsi Xu, Natalie Hutchinson, Thomas House).

## Abstract

The aim of this paper is to model homeless accommodation settings to investigate how vaccination mitigates the outbreaks, highlighting the importance of vaccination in vulnerable settings. We estimate the daily per capita contact rate with wider community, the internal transmission rate, and the achieved vaccine coverage. We present stochastic simulation of the final size of disease outbreaks given choices of internal and external transmission. We conclude that vaccine that has effect in reducing transmission will mitigate the outbreak in homeless hostels but it will have better results when the household population has large vaccination coverage, which may lead to more cost from the health economic perspective.

## 1. Introduction

In the UK, studies have indicated around 100,000 people cycle through the hostel system annually, many of whom have been previously rough sleeping (cf. Jones & Pleace (2010) and Story et al. (2014)). Sutherland et al. (2022) highlight that homeless populations frequently suffer from underlying health conditions such as diabetes, cardiovascular diseases, and respiratory problems. Concurrently, homeless populations have limited access to the primary care, which exacerbates the health problems. The study of Elwell-Sutton et al. (2017) shows that in England only 83.3% of single homeless in accommodation, 89.0% of hidden homeless people and 65.5% of rough sleepers were registered with a GP, which is much lower than the GP registration rate of English general population, 98.0%. Moreover, people currently experiencing homelessness may have additional difficulties in managing their exposure to potential infection, including social distancing, self-quarantine, and isolation Sutherland et al. (2022). These issues could compound and result in that the inpatient admission rate of homeless population is 3.2 higher than general community and the A&E (Accident & Emergency) attendance rate of homeless population is 5 times higher than that of general community (see Office of the Chief Analyst, Department of Health (2010)).

For influenza, there is evidence that homeless populations have particularly large spikes in hospitalisations during pandemic influenza seasons, suggesting vulnerability to viral respiratory infections Lewer et al. (2020). Moreover, the work of Story et al. (2014) highlights that 38.9% (95% CI: 31.5,48.2) of homeless individuals aged 16-64 have clinical risk factors for influenza, and lower than 25% (95% CI: 19.8,28.3) of these individuals received the vaccine, compared to over 53.2% in the general population. Boonyaratanakornkit et al. (2019) considers that homeless population is facing increased risk for poor outcomes from respiratory infections, indicating the important role of health interventions such as vaccinations. Influenza outbreaks in homeless populations can have broader public health implications, potentially spreading to the general population. Effective management and prevention of influenza in homeless communities can help reduce overall transmission rates and protect public health. This could be extended to other diseases and to other risk settings such as refugee camps Aylett-Bullock et al. (2022). The higher rates of hospitalization and emergency care among homeless individuals contribute to increased healthcare costs. Preventative measures, such as vaccination, can mitigate these costs by reducing the incidence of severe cases and hospitalizations.

Vaccination can mitigate disease outbreaks in three key ways: by reducing infection, lowering infectiousness, and decreasing disease severity. People that are vaccinated are less likely to become infected upon contact with infectious cases. This can be modelled two ways, as ‘all-or-nothing’ vaccine where there is a failure probability of vaccine but failure happens at dosage and successes are protected for duration of simulation, or as ‘leaky’ vaccination where failure happens at each contact opportunity. Furthermore, if people that are vaccinated are infected then the ability to transmit disease may be reduced. This can be modelled a parameter modifying infectious potential from breakthrough infections. From the perspective of reducing disease severity, people that are vaccinated suffer fewer complications and severe symptoms. This can be modelled as a scaling factor on cases and can be applied after simulation (thus assuming that severity of infection does not affect mixing behaviour). Given the uncertainty in the homeless setting we will not over complicate the modelling and instead assume that vaccine is ‘leaky’, reduces severity with a constant factor over all cases and has no impact on infectiousness (if vaccine fails to prevent infection then the case has a natural disease progression). The models described in Section 2 (and the Supplementary material) may allow greater flexibility and this may be investigated in future.

Given the potential role of vaccination in protecting homeless populations and reducing public health risk, our aim is to develop mathematical models to investigate how vaccination can mitigate the outbreak in homeless hostels and evaluate the cost and benefit of vaccine campaign from the economic perspective, aiming to provide support for policy making. In this report, we consider a scenario where influenza is introduced into a large ‘community’ population and through social interaction the influenza can be introduced to homeless settings with associated onwards transmission within setting. In this situation, a vaccination campaign is carried out to vaccinate people from both the community and the homeless population. We start from developing deterministic compartmental models to consider how vaccine protects people in a distinct subgroup and then extend it to incorporate the interaction between two populations, i.e. the household community and homeless hostels. Finally, we consider a stochastic ‘final size’ model and simulate outbreaks with and without vaccine in place. We compare different assumptions about vaccine effectiveness in the population at risk in reducing susceptibility or severity. In Section 3 the cost and benefit of the vaccine is evaluated and the impact is simulated in the homeless population.

## 2. Methods

### 2.1. Model overview and homeless accommodation structure

We developed a mathematical modelling framework to assess how seasonal influenza vaccination mitigates outbreaks in homeless accommodation settings. We considered a large community population experiencing a seasonal influenza wave, from which infections could be introduced into homeless settings through community contact and subsequently transmitted within settings.

The size and structure of the homeless population are difficult to estimate, not least because there are different types of homelessness. Each setting *i* will have a population *N*_*i*_ and we consider two types of accommodation: hostels and night shelters. Details of the data sources, accommodation-size distributions, and parameterisation of hostel and night-shelter populations used in this study are provided in Supplementary Material A.

### 2.2. Stochastic final size simulation

To assess impact in discrete locations, we introduce a Sellke construction simulator to consider the final size of an outbreak (cf. House et al. (2013)). We neglect the evolution over time because we have no calibration or validation data of outbreaks over time in settings so reporting total cases is sufficient for this study, though we include some temporal simulation results in Supplementary material B.

For each setting, infections could be introduced from the wider community and then generate onward transmission within the setting. The number of imported infections was sampled from a Poisson distribution with ingress rate Λ*N* where *N* is the number of residents/service users and Λ = *πλT* is the per capita contact rate. Here, *π* is the prevalence of influenza in the wider community, *λ* is the daily per capita contact rate between setting residents and the wider community, and *T* is the duration of the importation period. Because outbreaks in individual settings were assumed to last only a few weeks, community prevalence was treated as approximately constant over this period. The number of imported infections was capped at the setting population size.

A proportion *ω* of residents were assumed to be vaccinated before the season. Vaccination could reduce susceptibility to both imported and internally generated infections through the relative susceptibility parameter *ϵ* . Transmission within each setting was then simulated using the Sellke final-size construction, which is parameterised by the setting-specific reproduction number. The internal reproduction number in night shelters was denoted *R*_0,*S*_, and the corresponding reproduction number in hostels was *R*_0,*H*_ = ln(2)*R*_0,*S*_ . This means that the transmission in a hostel is 70% that of transmission within a night shelter, estimated to reflect the reduced contacts rates of individuals sleeping in lone rooms rather than shared dorms. This assumption would benefit from further research. Also, infections internal to setting are assumed to have a gamma distributed generation time with mean *µ* = 5 days and shape parameter *α* = 3 with reproduction number of *R*_0_ (the latter varied in simulations).

### 2.3. Parameterisation

Measuring the values of these parameters can be challenging due to the lack of effective surveillance to monitor respiratory viral infection trends in homeless settings. A literature review helped us identify values for key parameters. We varied the internal reproduction number in night shelters, *R*_0,*S*_, from 0 to 3, (meaning the corresponding reproduction number in hostels to *R*_0,*H*_ = ln(2)*R*_0,*S*_). These reproduction numbers in night shelters would translate, uncontrolled, to 80% attack ratios and in hostels to 50% attack ratios when *R*_0,*S*_ = 2 . Baseline vaccine coverage was set to *ω* = 0.73, as 73.2% (139/190), based on reported willingness to accept influenza vaccination among people experiencing homelessness (cf. Story et al. (2014)). Sensitivity analyses considered lower coverage values of 50% and 25%. Parameter values used in the stochastic final-size simulations are summarised in Table 1. The rationale for parameter choices and the assumed difference between hostels and night shelters is provided in Supplementary Material B.2.1.

**Table 1:** Parameter values used in the stochastic final-size simulations.

| Symbol | Description | Value & source |
| --- | --- | --- |
| $\Lambda$ | Import rate from community during outbreak | varied from 0.03 to 0.3 in steps of 0.03 in simulation |
| $\omega$ | Vaccine coverage | 0.73 (cf. Story et al. (2014)) |
| $\epsilon$ | Vaccine effectiveness at reducing susceptibility | 0.5 or 1 (no effect) (cf. Centers for Disease Control and Prevention (2023b)) |
| $\theta$ | vaccine effectiveness at reducing severity | 0.5 or 1 (no effect) (cf. Centers for Disease Control and Prevention (2023b)) |
| $R_{0,S}$ | Reproduction number in night shelters (internal) | varied from 0 to 3 in steps of 0.5 in simulations |
| $R_{0,H}$ | Reproduction number in hostels (internal) | $\ln 2 R_{0,S}$ (reduced due to reduced occupancy during sleeping) |

## 3. Cost-benefit analysis

We used a simplified cost-benefit framework to compare the expected cost of influenza outbreaks with and without vaccination in homeless accommodation settings. Let *C*_*U*_ denote the expected cost of an outbreak without vaccination, *C*_*V*_ the expected cost of an outbreak with vaccination, and *C*_*D*_ the cost of delivering the vaccination campaign. Vaccination was considered cost-beneficial when *C*_*D*_ *< C*_*U*_ − *C*_*V*_ .

### 3.1. Cost per infection

The mean cost per influenza infection, *µ*_*C*_, included quality-adjusted life year (QALY) losses, primary care costs, hospitalisation costs, and mortality costs. It was calculated as

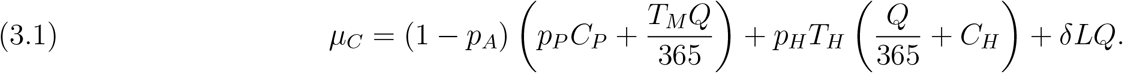

in which *p*_*A*_ is the probability that infection is asymptomatic; *p*_*P*_ is the probability that a symptomatic case seeks primary care; *C*_*P*_ is the cost of a primary care consultation; *T*_*M*_ is the duration of mild illness; *Q* is the monetary value of one QALY; *p*_*H*_ is the probability of hospitalisation; *T*_*H*_ is the duration of hospitalisation; *C*_*H*_ is the daily hospital cost; *δ* is the mortality risk; and *L* is the number of life years lost per influenza death. Baseline values are shown in Table 2, giving *µ*_*C*_ = 461.19 (£) per infection. More detailed explanation for these parameter values is provided in Supplementary Material C.

**Table 2:** Parameters for cost-benefit analysis

| Symbol | Description | Value & source |
| --- | --- | --- |
| $T_M$ | Duration of mild illness | 3.8 (days) |
| $T_H$ | Duration of hospitalisation | 8 (days) |
| $L$ | Life years lost by premature death | 10 (years) |
| $C_P$ | Cost of primary care | 56 (£) |
| $C_H$ | Cost of secondary (hospital) care per day | 350 (£) |
| $Q$ | Societal QALY | 20000 (£) |
| $p_A$ | Probability of case being asymptomatic | 0.4 |
| $p_P$ | Probability of seeking Primary Healthcare | 0.2 |
| $p_H$ | Probability of hospitalisation | 0.04 |
| $\delta$ | Probability of death | 0.001 |
| $((1 - p_A)T_M + p_H T_H)/365 + \delta L$ | Scaling factor on QALY | 0.0171 |
| $(1 - p_A)p_P C_P + p_H T_H C_H$ | Non-QALY costs | 118 (£) |
| $\mu_C$ | Total average cost of case | 461.19 (£) |

### 3.2. Vaccination campaign cost

We assume campaign costs are evaluated for a single season, so there is no discounting of accrued cost into the future, and include purchasing vaccines, storage and deployment to settings and physical delivery to individuals by healthcare workers. The cost of delivering vaccination in a setting of size *N* was calculated by:

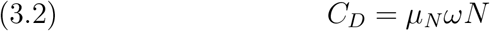

where *µ*_*N*_ is the cost per vaccinated individual and *ω* is the vaccine coverage.

Vaccination cost assumptions are summarised in Table 3. Under the baseline assumptions, the cost per vaccinated individual was

**Table 3:**
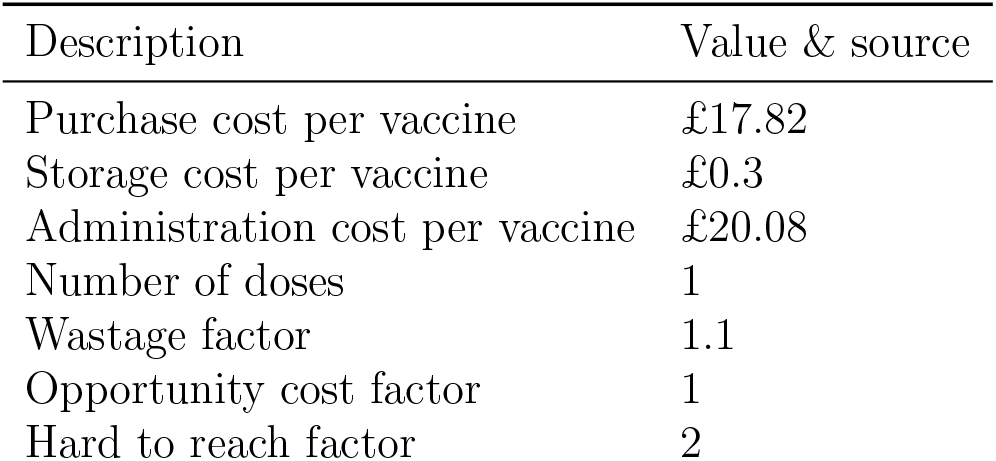
Cost components and scaling factors used to calculate the vaccination campaign cost.

| Description | Value & source |
| --- | --- |
| Purchase cost per vaccine | £17.82 |
| Storage cost per vaccine | £0.3 |
| Administration cost per vaccine | £20.08 |
| Number of doses | 1 |
| Wastage factor | 1.1 |
| Opportunity cost factor | 1 |
| Hard to reach factor | 2 |

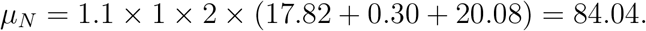

in which 1.1 is the wastage factor, 1 is the opportunity cost factor, and 2 is the hard-to-reach delivery factor. The hard-to-reach factor reflects the assumption that approximately half of vaccinated individuals would receive vaccination through standard primary care, while half would be reached through targeted outreach services at three times the standard delivery cost. This gives an average delivery multiplier of (0.5 × 1) + (0.5 × 3) = 2 .

Therefore a campaign covering 73% of all hostel and shelter service users (approx 40,000) would be about 2.4 million. Note that analysis of programmatic data from Find and Treat commissioned within UKHSA suggested a lower vaccination cost of approximately £54 per person, including vaccine, delivery, and wider programme costs. We retained £84.04 as the baseline value to avoid underestimating campaign costs.

### 3.3. Integration with stochastic final-size simulations

For each stochastic final-size simulation, let *X*_*U*_ and *X*_*V*_ denote the final number of infections in the settings without and with vaccination respectively. The outbreak cost without vaccination was calculated as:

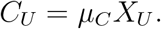

And the outbreak cost with vaccination was approximated as

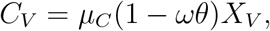

where *θ* is the proportional reduction in disease burden among vaccinated infected individuals. This formulation allows vaccination to reduce costs through two mechanisms: by reducing the number of infections through susceptibility reduction, and by reducing the clinical and economic burden of infections occurring after vaccination. Furthermore, for a setting of size *N*, the vaccination campaign cost was

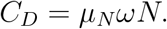

As we mentioned above, vaccination was considered cost-beneficial when *C*_*D*_ *< C*_*U*_ − *C*_*V*_, which is equivalent to

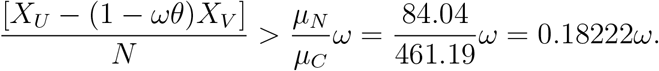

This framing is intuitive from a disease outbreak viewpoint. Note that we can define the notation *R*_*∞*_ = *X*_*U*_ */N* as the attack ratio in a setting (expected cases divided by total population) pre-vaccination (at baseline) and similarly define 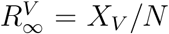. In this notation we can state 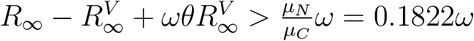.

So assuming no impact on susceptibility 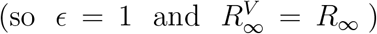, then the vaccine coverage cancels on both sides and this calculation simplifies to 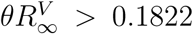. So with *θ* = 0.5 then vaccine is beneficial provided the unvaccinated attack ratio *R*_*∞*_ *>* 0.3644 . This could be a reasonable rule of thumb (i.e. a general indication of vaccine benefit in the homeless population would be if in absence of vaccine campaign one observes 15000 cases per season, noting that many of these may be asymptomatic.). In a stochastic simulation we simulate the number of cases in an outbreak (rather than proportion of cases) so we apply these outbreak costs by individual rather than these population averages to get greater variation to results. Noting that the less risk adverse cost per case is £900 rather than £460 this would half the threshold attack ratio rule of thumb. Moreover using the value of £54 for campaign cost per dose (with the higher cost per case) would suggest a much lower threshold attack ratio of 12% (given 50% severity reducing effectiveness of vaccine).

## 4. Simulation Results

We treat the costs of vaccine campaign and the cost of disease episodes as fixed (though allow stochastic effects through sampling the duration of illness and probability of severe outcomes as Poisson and binomial events respectively).

The economic evaluation in the previous section suggests that the cost of a case is about 5.5 times that of a vaccination (£461.19 versus £84.04). Remember we have deliberately chosen lower value for some costs associated with cases. This means if in a setting we expect 20 cases in a season without vaccine (£9,224.4) and this setting has 50 residents of which 75% accept vaccine the campaign would cost £3,151.5. Thus, the vaccination campaign would need to avert approximately 7 equivalent infections to be cost effective. Doubling the vaccine costs would approximately double this threshold.

### 4.1. Single Stochastic Outbreak Simulation

Figure 1 shows a single stochastic simulation of the costs presented above. Here 30 people suffer infection represented by each increment on *x* -axis. The *y* -axis is the time period people are ill for, with the equivalent financial impact written above each bar. Yellow bars show mild illness (seen in 16 out of 30) whilst red bars show periods in hospital (in this case only seen in 2 cases). The cost of the unvaccinated outbreak is then given by the sum of the values above each bar (£14,026.86). We assume vaccine coverage of 73% and those individuals receiving *effective* vaccine (vaccine effectiveness against severity is 50%) are marked with blue dot at base of each bar (and so the total cost associated with those bars are removed from outbreak cost given the vaccine campaign - on the basis that those vaccinated cases would avoid the impact of disease). If the vaccine was susceptibility reducing then the outbreak would have reduced in this simulation from 30 to 6 cases (those cases marked with a purple dot).

**Figure 1:**
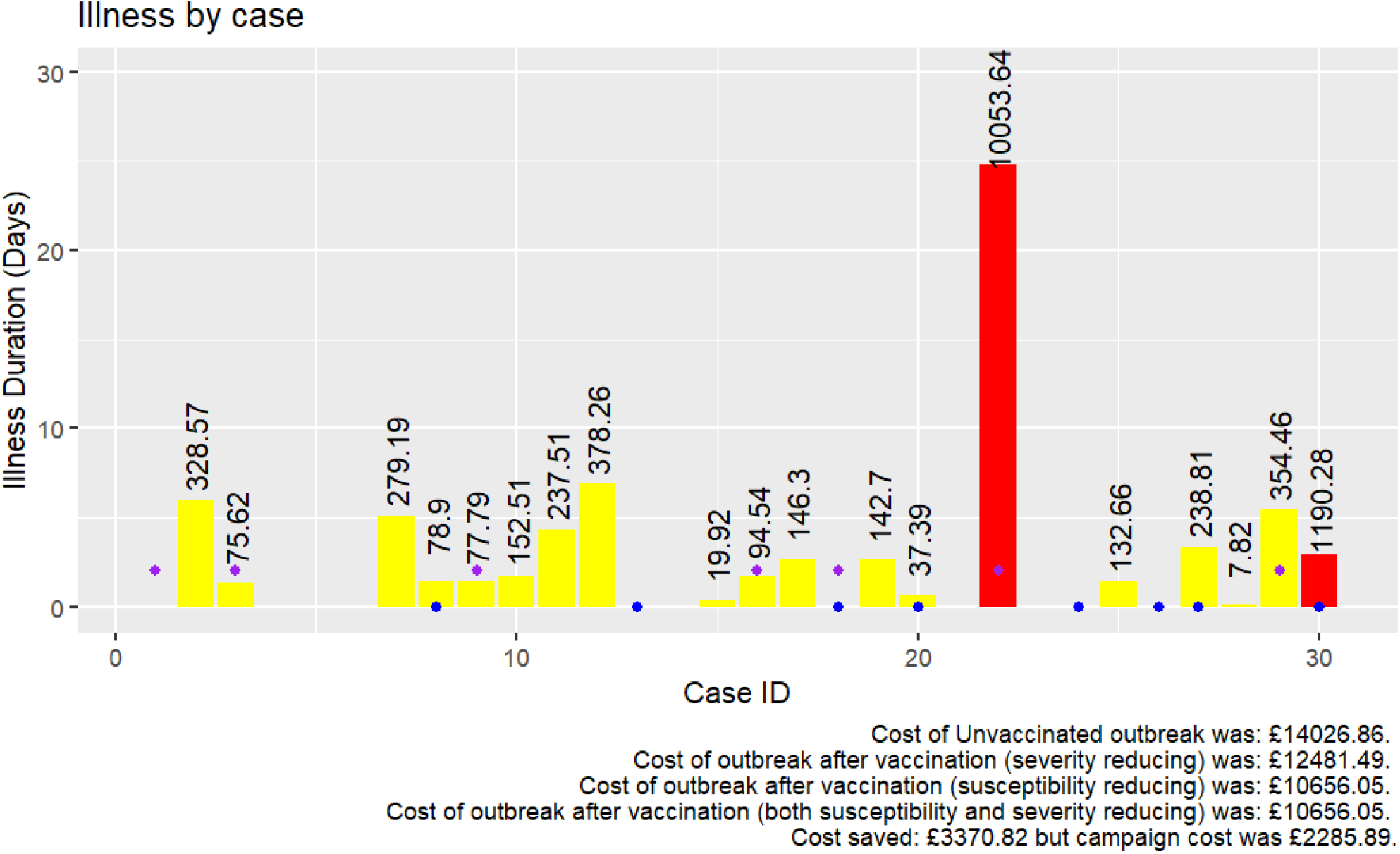
Single stochastic simulation of the cost per individual. Here 30 people suffer infection. The y-axis is the time period people are ill for, with the equivalent financial impact written above each bar. Yellow bars show mild illness whilst red bars show periods in hospital. Blue dots represent cases avoided due to severity reduction from vaccination and purple dots represent cases arising from post-vaccine outbreak where vaccine is susceptibility reducing.

Despite this simulation being ‘unlucky’ in terms that the hospitalised case with longest length of stay is simulated to occur even after vaccination and so the outbreak costs remain high there is still a saving of £3,370.82 in terms of disease impacts averted. This is in excess of the cost of the campaign itself of £2285.89. In present simulation the susceptibility and severity sampling is done independently which is credible if vaccine coverage is 100% but further refinement will investigate impact of this for lower coverage values.

### 4.2. Ensemble stochastic results

Figure 2 shows the results of simulating a season of influenza in all 1208 homeless settings (143 night shelters, 1065 hostels). Each panel represents different assumptions on vaccine effectiveness (left panel severity reducing only, middle panel susceptibility reducing only and right panel combined effectiveness) with changes to community force of infection per user onto setting (*λπT*) and internal transmission (*R*_0,*S*_).

**Figure 2:**
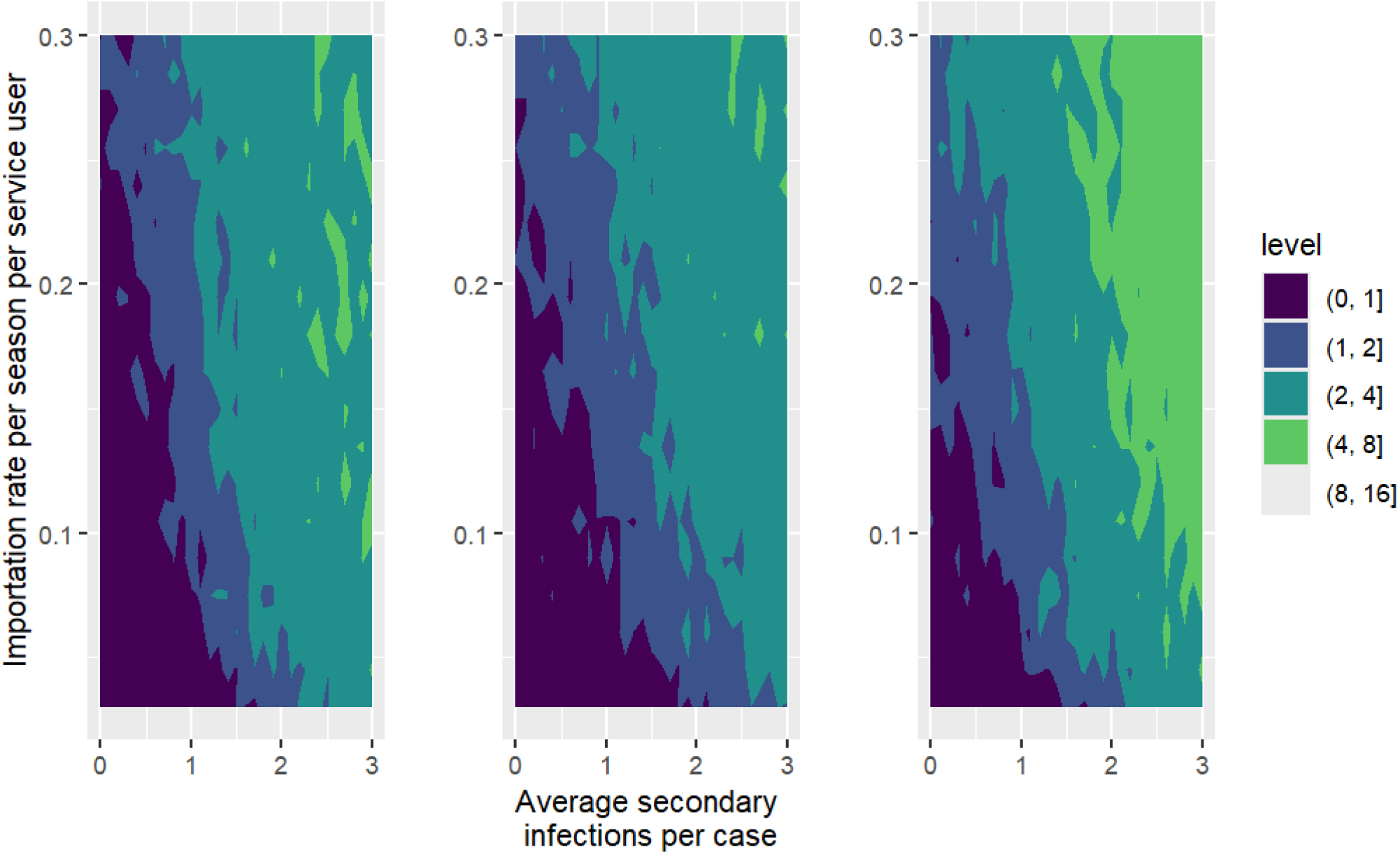
Mean cost effectiveness arising from simulation of outbreaks in 1208 homeless settings assuming vaccine coverage of 73% assuming vaccine is severity reducing only (left panel), susceptibility reducing (middle panel) and both (right panel). The *x* -axis in each panel is the control reproduction number in setting and the *y* -axis the ingress rate of cases from community. The levels marked by contours show the relative mean cost (total cost of cases averted in outbreaks given vaccination divided by total cost of campaign). Those parameter combinations that were not cost effective are marked in darkest colour (levels less than 1 on legend scale).

Within each panel the *y* -axis shows ingress from community increases (*λ* from 0.1 to 1 in steps of 0.1) this means the overall ingress rate ranges from 0.03-0.3 (meaning a setting of 33 people would experience ingresses range from about 1 to 10 per season). The *x* -axis shows the control reproduction number (average number of secondary infections in otherwise susceptible population at risk) for values from *R*_0,*S*_ = 0 up to *R*_0,*S*_ = 3 .

With no internal transmission the ingress rate must be very strong to show benefit to vaccine. If internal transmission is strong (greater than 2) vaccine appears beneficial (as might be expected from basic theory as it would be likely to observe *>* 80% attack ratios in settings if transmission was that strong). For moderate to low internal transmission vaccine can be effective but depends on the ingress rate.

The mean result may be skewed to larger settings (particularly this may be an issue given the tail in simulated setting sizes). Figure 3 has the same panel ordering but shows the proportion (quantile in level incrementing in 10%) of simulations where the simulation was cost effective (above the relative cost threshold of 1). This is less sensitive to larger settings but to interpret one must chose a decile for decision making. This may be 50% (so that half the settings are better off). As with Figure 2 this shows that if transmission is expected to be strong in homeless setting then vaccine is cost effective. So better understanding of likely secondary transmission in the settings are critical. Similar plots for different values of vaccine coverage (25% and 50%) are shown in Supplementary Material D.

**Figure 3:**
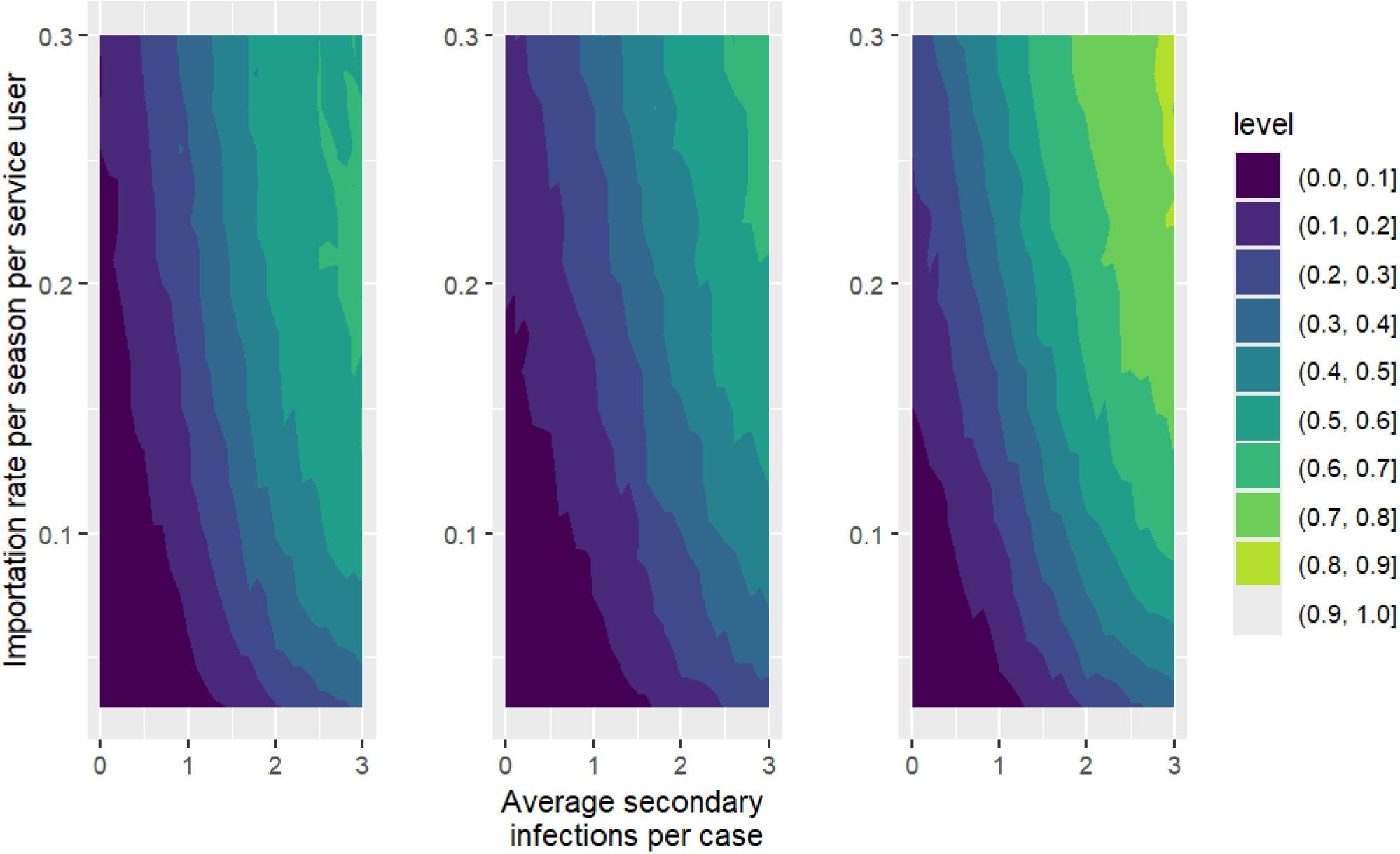
Sample quantile of the simulations that were cost effective for 1208 homeless settings assuming vaccine coverage of 73% assuming vaccine is severity reducing only (left panel), susceptibility reducing (middle panel) and both (right panel). The *x* -axis in each panel is the control reproduction number in setting and the *y* -axis the ingress rate of cases from community, the levels marked by contours show the proportion of simulations that were cost effective

Instead we can consider total cases arising from an internal and external transmission scenario. Figure 4 shows an expected result (see above) that a severity only reducing vaccine would be cost effective if around 15,000 cases arose in seasons without vaccination and is independent of coverage (black points similar subject to random noise in each panel). However, susceptibility reduction has a strong effect, even when not coupled with severity reduction though this impact reduces as coverages increases.

**Figure 4:**
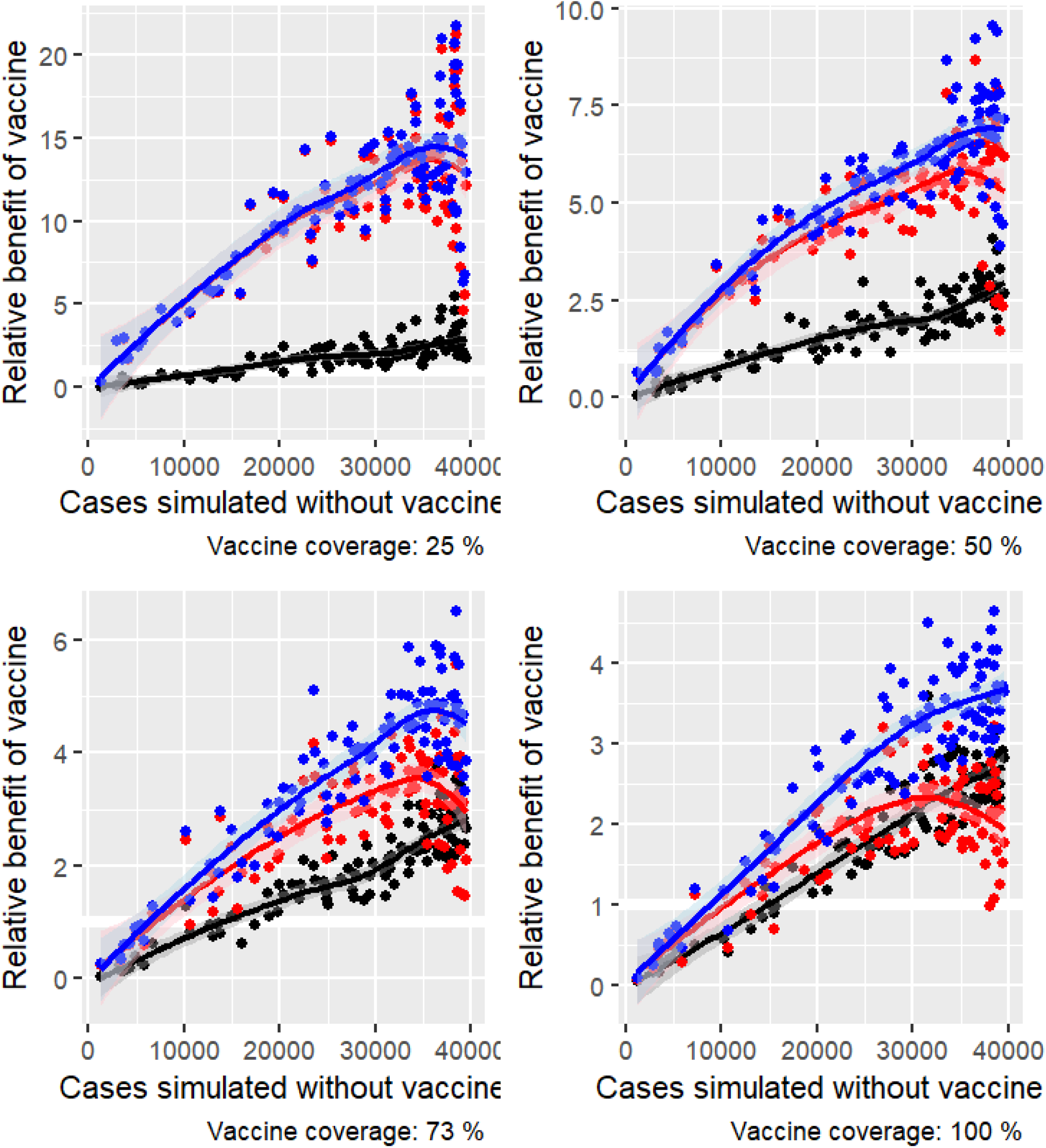
Figures showing the relative benefit of vaccine campaign given cases expected prior to vaccination for different coverage rates in population. Black line in each panel represents assuming severity-reducing-only vaccine, red is susceptibility-reducing-only vaccine and blue would be both susceptibility and severity reduction.

There are perhaps two counter-intuitive features in the results in Figure 4. Firstly for 73% coverage (bottom left) the vaccine is cost effective if around 5,000 cases are observed without vaccine, and this threshold reduces as coverage drops. We might expect lower coverage rates to be worse rather than better but this is due to the associated campaign costing less. This could tell us the coverage to aim for assuming a given number of cases pre-vaccination if surveillance data was available.

Secondly at higher attack ratios in the at risk population the results exhibit greater variation when vaccine is susceptibility reducing. So for the same number of total cases arising from unvaccinated outbreaks the benefit of vaccine varies according to the balance between internal and external transmission generating those case numbers. Partly this is because in simulation external ingress acts first so if ingress rate is high (expectation of an infection from outside is about 1 or higher) then the vaccine effect acts on all cases. If ingress is more moderate (so we expect all settings to suffer an ingress of disease to cause an outbreak but the outbreak is then sustained by internal transmission) this will be subject to greater variation.

Consider 40,000 cases pre-vaccination (roughly the total homeless population that use the services). If these all come from external infection from local community and a campaign has complete coverage with 50% efficacy then both susceptibility or severity reducing vaccine would generate 20000 cases because the community outbreak is not benefiting from vaccine (susceptibility and severity reducing would generate 10,000 cases). If ingress is lower then internal transmission can benefit from susceptibility reduction and reduce attack rates. However, if *R*_0_ = 4 pre-vaccine we would expect a 99% attack ratio, but susceptibility reduction would lead to *R*_*V ACC*_ = 2, leading to an 80% attack ratio and so we would expect 32000 cases (8,000 cases averted). This is unlikely to be an issue in reality as attack ratios would have to be very high and require the vaccine to only be susceptibility reducing.

## 5. Discussion

This work should be viewed as an initial scoping of the challenges of modelling homeless setting outbreaks and so indicative rather than definitive results. Further work is needed to refine assumptions and verify model structures to improve future disease impact predictions. It is our view that reporting the struggles to generate data-driven evidence based simulations is itself important.

Modelling disease outbreaks in homeless settings is highly uncertain. The major parameteric uncertainties are the daily per capita contact rate with wider community, the internal transmission rate (parameterised here in terms of a ‘reproduction number’) and the achieved vaccine coverage.

The lack of enhanced surveillance in such settings for respiratory viral infection mean that the number of infections generated within the setting through local contacts is unclear.

There are a number of limitations or assumptions, and so we present this work as a starting point for further work. We have assumed the vaccine is leaky in reducing susceptibility but that it is all or nothing in terms of severity reduction. So in Figure 1 any case that had received effective vaccination (blue dot) has all their disease impact reduced. We could have instead assume that severity was reduced proportionately (effectively shortening their illness by factor of *θ*). This is equivalent to assuming severity reduction is all-or-nothing rather than leaky. This is not likely to impact the overall average conclusions (in the way assuming susceptibility reduction is leaky does), but likely increases the variation in the results from simulation. Assuming an impact on infectivity from vaccinated cases will improve the benefit of vaccination and so these results are in that sense pessimistic.

The disease progression parameter assumptions are uncertain but based on studies from real world, albeit not all from similar settings. Conducting enhanced surveillance in homeless shelters and hostels would be sensible, to help assess disease impacts on individuals but also the difference in transmission between setting types since the current model assumes that transmission in hostels is around 70% of that in night shelters.

However, note that the baseline cost per infection is substantially driven by QALY-related losses. Mortality contributes around £200 of the £461 total cost per infection, while hospitalisation and mild illness each contribute approximately £130. If the case fatality ratio or life years lost were lower, the benefits of vaccination would be overestimated; improved assessment of frailty and influenza impact in this population is therefore critical.

Given the uncertainty in the parameter values and modelling assumptions we have taken a ‘big picture’ approach to presenting the results. Clearly, if better characterization of the transmission and ingress terms (and also generalizable to homeless accommodation estate) were available, greater sensitivity could be conducted on vaccine coverage and efficacy values that may provide edge cases in decision space.

## Data Availability

All model input parameters used in this study were derived from published literature and publicly available reports cited in the manuscript and supplementary material. This study did not use individual-level data, patient records, or restricted-access datasets. Model outputs generated in the present study are included in the manuscript and supplementary material.

## A. Supplementary Material: Accommodation Structure and Setting-size Distribution

The size and structure of the homeless population are difficult to estimate, not least because there are different types of homelessness. People who live in hostels for single homeless individuals typically reside in single-bedroom accommodation. A census of accommodation providers for single homeless people suggested that 35,817 people lived in 1,065 hostels in England in 2019, and the median size of the hostel was 21 beds (IQR 12–38) Lewer et al. (2020). In the absence of further data, we assume that the hostel size *N*_*H*_ is a random variable with a shifted Negative Binomial distribution *N*_*H*_ ∼ *M* +NB(*n*; *µ* − *M, r*) (in this case the minimum size of *M* = 12 beds, mean of *µ* = 33.63 beds, and size parameter *r* = 0.4 gives a median of 20 beds (IQR 13-39)).

Few estimates of the number of people sleeping rough are available. An official government count in combination with an estimate of the number of people not captured by this count, based on data from a multi-agency database, gives an estimate of 10,748 people currently sleeping rough in England Lewer et al. (2020).

For night shelters, which means multiple beds in one large room, we assumed that people sleeping in night shelters are a subset of this rough sleeping population. Data on the number and size of night shelters in London was used to estimate that 3,616 of 10,748 people sleeping rough in England are sleeping in 143 night shelters Lewer et al. (2020). These night shelters have a median of 15 beds (IQR 14–25) each Lewer et al. (2020). As above we assume that the shelter size is random variable with shifted Negative Binomial distribution (minimum size of 14 beds, mean of 25.29 beds and size parameter 0.22).

These inferred negative binomial distributions capture the lower tail of empirically reported sizes but due to the size parameter being less than 1 we note that some simulated settings have unrealistically large simulated populations. The violin plots in Figure 5 show the distribution of residents simulated from the information provided (left hand hostels, right hand night shelters). The green horizontal solid line shows the median number of people per setting, the green vertical line the upper and lower quartiles stated in Lewer et al. (2020). The bulk of the simulated distribution is therefore plausible with evidence. However the extreme upper tail is potentially high, better characterization of the size of hostels and shelters would be critical in future.

**Figure 5:**
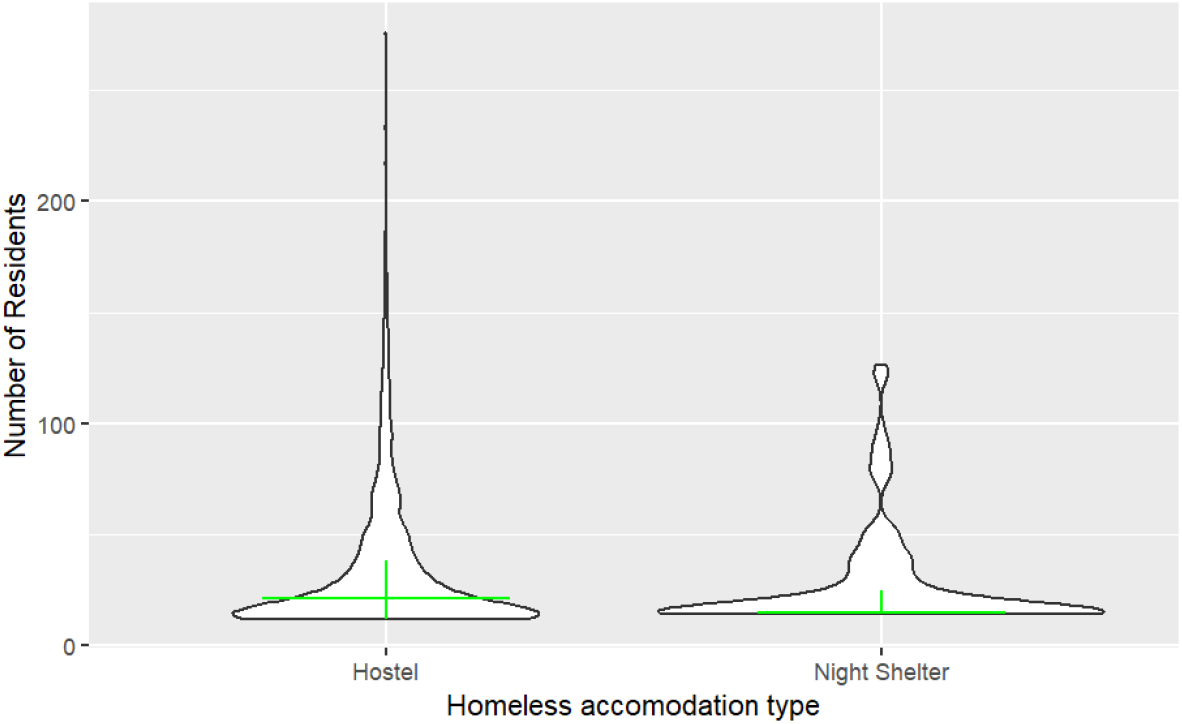
The simulated distribution of residents derived from Lewer et al. (2020) (left hand hostels, right hand night shelters). The green horizontal solid line shows the median number of people per setting, the green vertical line the upper and lower quartiles

## B. Supplementary material: A temporal model

### B.1. Disease and vaccination assumptions in modelling

We assumed that no outbreak was present in homeless accommodation settings at the start of the season. Vaccination was assumed to occur before the seasonal influenza wave, with a proportion *ω* of residents vaccinated at *t* = 0 . The initial conditions for a setting of size *N* were:

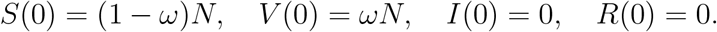

We could allow some natural initial cross-immunity by setting *R*(0) /= 0 . Such prior residual immunity will be hard to measure and so this is not investigated, but as a result this may mean that simulations are over estimates of potential effects.

For a single setting, the transmission dynamics were given by:

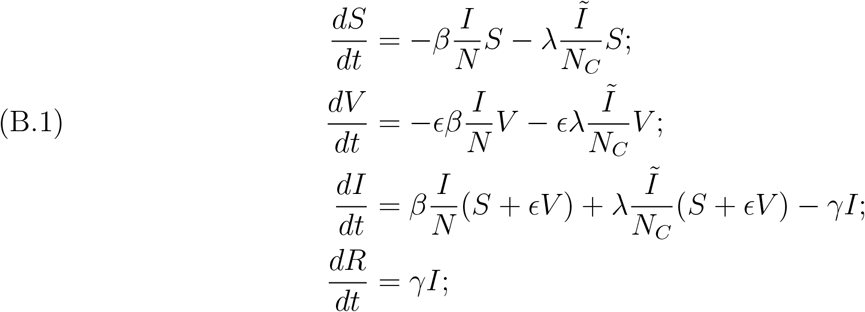

in which *S, V, I*, and *R* denote susceptible, vaccinated, infectious, and recovered individuals in the setting; *β* is the internal transmission rate; *γ* is the recovery rate; *λ* is the daily contact rate with the wider community; and 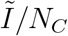 is the prevalence of infection in the community. Whilst amplification from a setting to the community can occur given the focus here is on the impact of vaccination on the setting population itself we neglect the interaction between homeless population back onto the community. Also, we did not include waning immunity or reinfection within a single influenza season; individuals who recovered from infection were assumed to remain immune for the duration of the season. Disease severity was assumed not to directly affect transmission behaviour. Instead, vaccination was allowed to reduce susceptibility to infection, through the parameter *ϵ*, and/or to reduce the clinical and economic burden of infection after infections were simulated, through the parameter *θ* . We assumed no reduction in infectiousness among vaccinated breakthrough infections.

### B.2. Compartmental Model for Population

To illustrate a model of vaccination we take a standard SIR model. Consider a large community of constant size *N*, where individuals can be in states, *S*, susceptible to infection, this population may get infected and so enter state *I*, from which they will get recovered after a certain period of time and join the state *R* . This model is intended to be illustrative and so we do not include further complexity in the infection dynamics although we expect factors such as waning immunity (and people who recover from the infection will have permanent immunity), or incubation period in reality. We do not at this stage consider the severity of the disease on infected and recovered cases. A vaccination programme to mitigate the outbreak is started. Hence, the population may get vaccinated, entering a state *V* . However, this vaccinated population can still get infected, *I*_*V*_ . We assume that vaccination does not affect the timing of recovery (but may affect the severity of infection). These dynamics is described by the following equations:

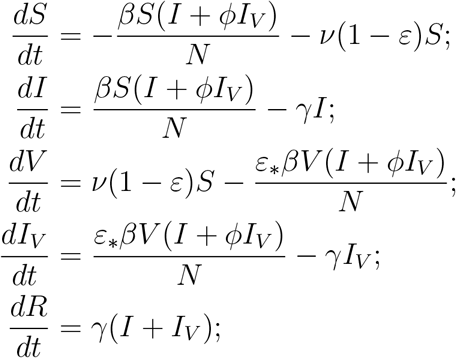

where *β* is the infection rate, *γ* the recovery rate, *ε* is the failure rate of the vaccine (1-efficacy or 1-effectiveness) (assuming everyone in population is offered vaccine, if there is a stockpile issue or other coverage impact then the interpretation of *ε* would change) and *ϕ* the reduction in transmission potential of vaccinated cases. Then *ε*_*\**_ is the vaccine effectiveness in reducing infection at each contact (a leaky vaccine where effect is per contact rather than all or nothing modelled by *ε*). Reduction in symptoms/severity can be a scaling factor on *R* but is assumed not to affect the transmission dynamics. Note that in this model the susceptible population would be continually vaccinated and eventually approach zero so the long term dynamics are likely flawed. In reality there maybe hard to reach groups or people that refuse vaccine that mean high coverage is not achievable.

This model can be further simplified by assuming vaccination occurs *prior* to introduction of disease (or *ν* is very fast). Then we can set *ν* = 0 and treat vaccination as initial condition (so *S*(0) = *N* − *ω*(1 − *ε*)*N* − *I*_0_, *I*(0) = 1_0_, *V* (0) = *ω*(1 − *ε*)*N* then *R*(0) = *R*_*V*_ (0) = *I*_*V*_ (0) = 0). Here *ω* is the fraction of the population at risk that accept the vaccine offer.

In the following subsections, we will pursue analytical work for different scenarios. Clearly these are perverse extreme limits but are illustrative of potential impacts from more structured and carefully parameterised models. We also define *β* = *ργ* so that the infection rate is described in terms of the recovery rate and a non-dimensional factor.

#### B.2.1. Transmission parameter assumptions

Measuring the values of these parameters can be challenging due to the lack of effective surveillance to monitor respiratory viral infection trends in homeless settings. Many studies consider an indicative mean value of *R*_0_ of influenza in the community to be 1.2, *R*_0_ ∈ [1.1, 1.5], across individuals (cf. Cowling et al. (2010), Opatowski et al. (2011), Roberts & Nishiura (2011)). The work of Brugger & Althaus (2020) measures the maximal *R*_0_ to be 1.46 to 1.81 (median). Assessing the reproduction number and porting to another setting is incredibly challenging as this is a combination of viral, behavioural and societal factors that change by location and setting. A literature review Finnie et al. (2014) showed that enclosed setting have higher attack ratios, while homeless settings are not truly enclosed as people are free to leave or move between such settings this serves is an indication that proximity and density elevate transmission.

Transmission is assumed to be different in night shelters and hostels due to the difference in accommodation. We assume the types of contact are similar and so the probability of transmission is the same in both types of setting. It is then the number of contacts that increases between hostel and night shelter. There is very little data to make informed choice on the difference in risk and this should be a future research question. Here we assume that contacts in hostels are around ln 2 ∼ 70 % than those in night shelters.

Another important parameter is the recovery rate. Influenza recovery rate has been estimated to be 3.8 days with 95% confidence interval (CI) 3.1-4.6 days McCarthy et al. (2020) (also cf. Cauchemez et al. (2004)). Hence we will adopt that the influenza recovery rate is 1/3.8 per day. We set vaccine coverage baseline to be *ω* = 0.73, as 73.2% (139/190), of homeless population in study shows willingness to take influenza vaccine if they were offered Story et al. (2014). We test eventual impact of this assumptions with sensitivity to values of 50% and 25%.

#### B.2.2. Special cases: no effect on reducing transmission

Here, we assume that the vaccination will have no effect on reducing transmission, i.e. *ϕ* = 1 . In this situation, we can define *Î* = *I* + *I*_*V*_ so that

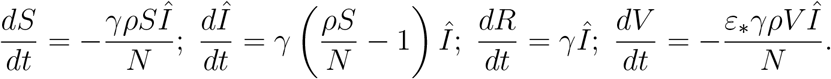

We can also simply get expression for final attack ratio *R*_*∞*_ = *R/N*, namely note that we can write

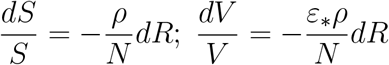

so *S* = *S*(0)*e*^*−ρR/N*^, 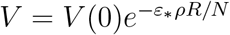 and remember that *S* + *V* + *Î*+ *R* = *N* so

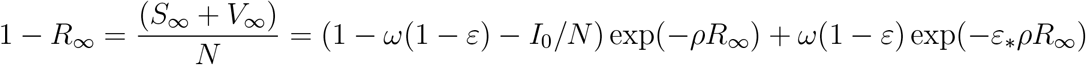

For illustration if *ω* = 1, *I*_0_ ≪ *N, ρ* = 2 (so uncontrolled we may expect an 80% attack ratio) and with vaccine we observe *R*_*∞*_ = 1*/*2 then we find *ε* and *ε*_*\**_ related with the following

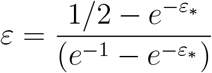

so if *ε* = 0 then *ε*_*\**_ = ln(2) . If *ε*_*\**_ = 0 then *ε* = (2(1 − *e*^*−*1^))^*−*1^ .

The assumption on all or nothing or leaky vaccination does not change the early growth rate (the ODE for *Î* has no *ε* factor) but does affect the final size. This is because in the leaky case you are affecting the whole population whilst in the all or nothing a subset are unprotected but unaware of this.

At present in the Homeless setting modelling we assume that *ϕ* = 1 (no impact of vaccine on transmission potential of vaccinated cases) and that *ε* = 0 (so vaccine is leaky and does not confer all-or-nothing status). The benefit of vaccine is then through modifying chance of infection at each contact and reducing severity. We investigate briefly the theoretical impact of transmission reduction in next section.

#### B.2.3. Different levels of effectiveness in reducing transmission

We consider the different levels of vaccine effectiveness in reducing transmission, i.e. different values of *ϕ* . When *ϕ* = 1, the vaccine has no effect in blocking transmission and when *ϕ* = 0, the vaccine will fully stop the transmission. To simplify the simulation, we assume that *ϵ* = 1, and *ν* = 0.002 . In Figure (6), we vary the value of *ϕ*, and we can see that:

**Figure 6:**
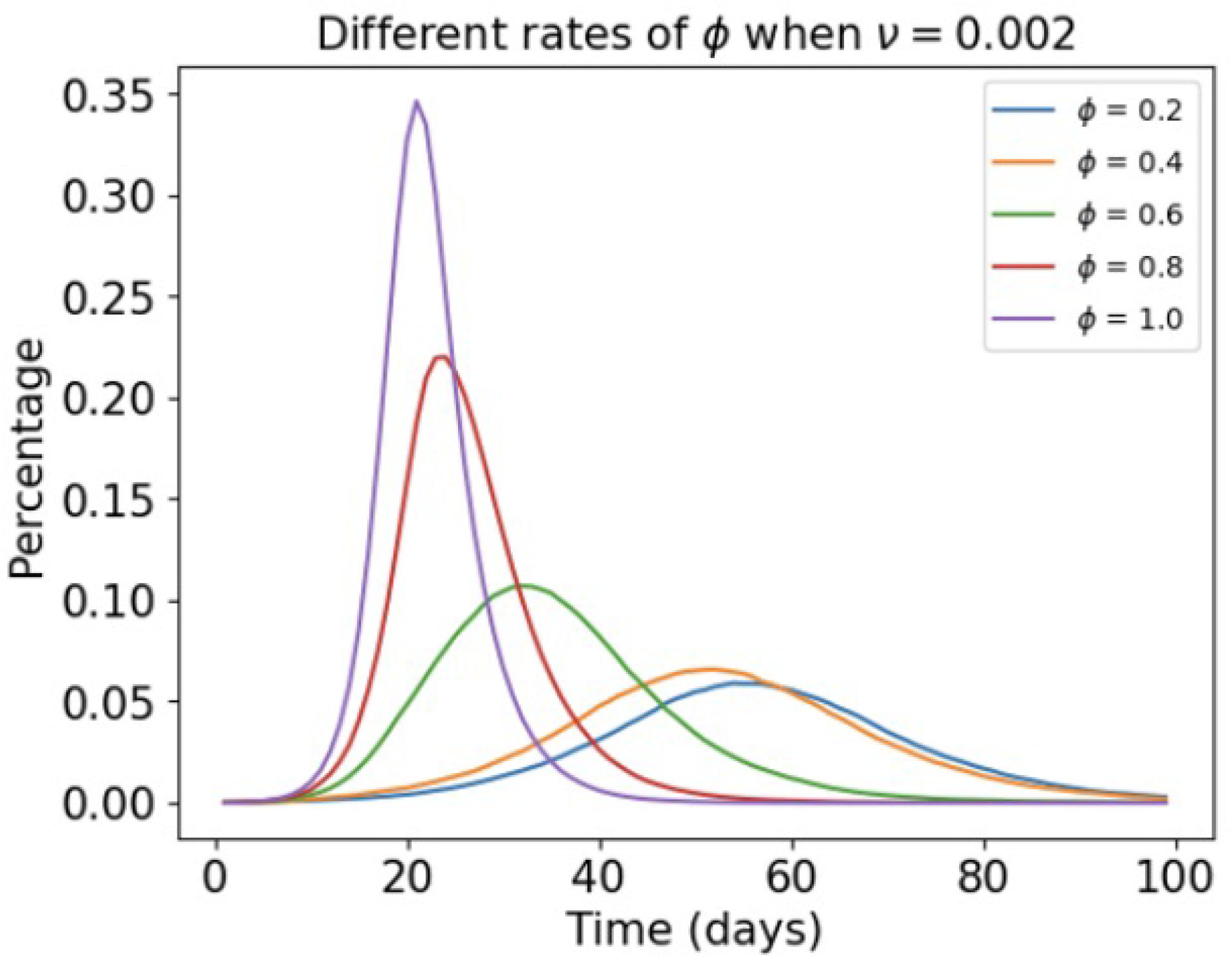
Figure showing the percentage of infected cases in the homeless population over time (in days) under different values of *ϕ* . Note that we set *ν* = 0.002 during the simulation.

- Intuitively, smaller value of *ϕ* will lead to better result in mitigating outbreak in homeless hostels. However, compared with the previous scenario, small values such as *ϕ* =
- 0.2 or 0.4 cannot fully control the outbreak. The reason is the low vaccination coverage in household population. In this situation, there is a large portion of non-vaccinated infected population, which leads to significant external infection force and vaccine mainly reduces the in-hostel transmission.
- In Figure (7), we increase the vaccine rate *ν* from 0.002 to 0.01, which means that the household community will have a large vaccination coverage. In this situation, we can see that reducing the value of *ϕ* will have a much better result in controlling the outbreak in homeless hostels.
- We can summarise that vaccine that has effect in reducing transmission will mitigate the outbreak in homeless hostels but it will have better results when the household population has large vaccination coverage, which may lead to more cost from the economic perspective.

**Figure 7:**
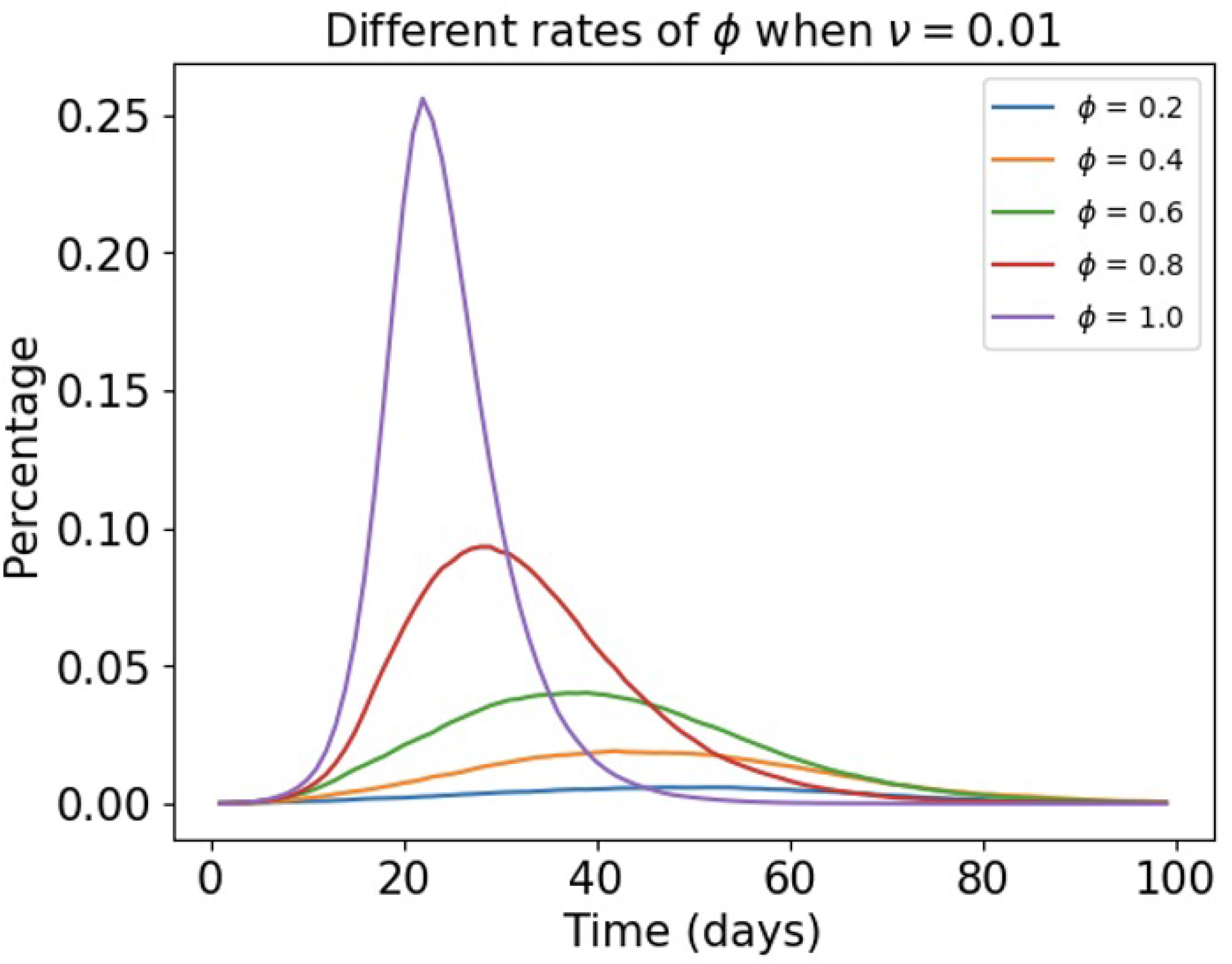
Figure showing the percentage of infected cases in the homeless population over time (in days) under different values of *ϕ* . Note that we set *ν* = 0.01 during the simulation.

#### B.2.4. Setting to Community model: Different levels of effectiveness in reduction in susceptibility

Given the lack of surveillance data from such settings we are not able to calibrate to a timeseries of case numbers and so instead can consider the final size of outbreaks.

During the simulation, we assume that there is no between hostels infection, which means it is a independent household model. We set there are 10,000 hostels and each hostels are independent. The simulation runs for 100 days and daily infection rate is illustrated in the following figures. With the parameters above, we set the infection rate to be *β* = 0.4 per day.

We carry out sensitive analysis to see how different parameters influences the outbreak in homeless hostels.

Here, we focus on the vaccine effectiveness in reduction in susceptibility, i.e. changing the value of *ϵ* . Smaller values of *ϵ* indicate stronger results in protecting people. Where *ϵ* = 1, the vaccination has no effect in reducing susceptibility and when *ϵ* = 0, the vaccine will completely protect the person. For the rest parameters, we set *ϕ* = 1, which means there is no reduction in transmission, and *ν* = 0.02, indicating that the community population is slowly getting vaccinated. We set *λ* = 0.3 which reflects the low interaction between homeless population with general population. In Figure (8), we vary the value of *ϵ*, and we notice that:

**Figure 8:**
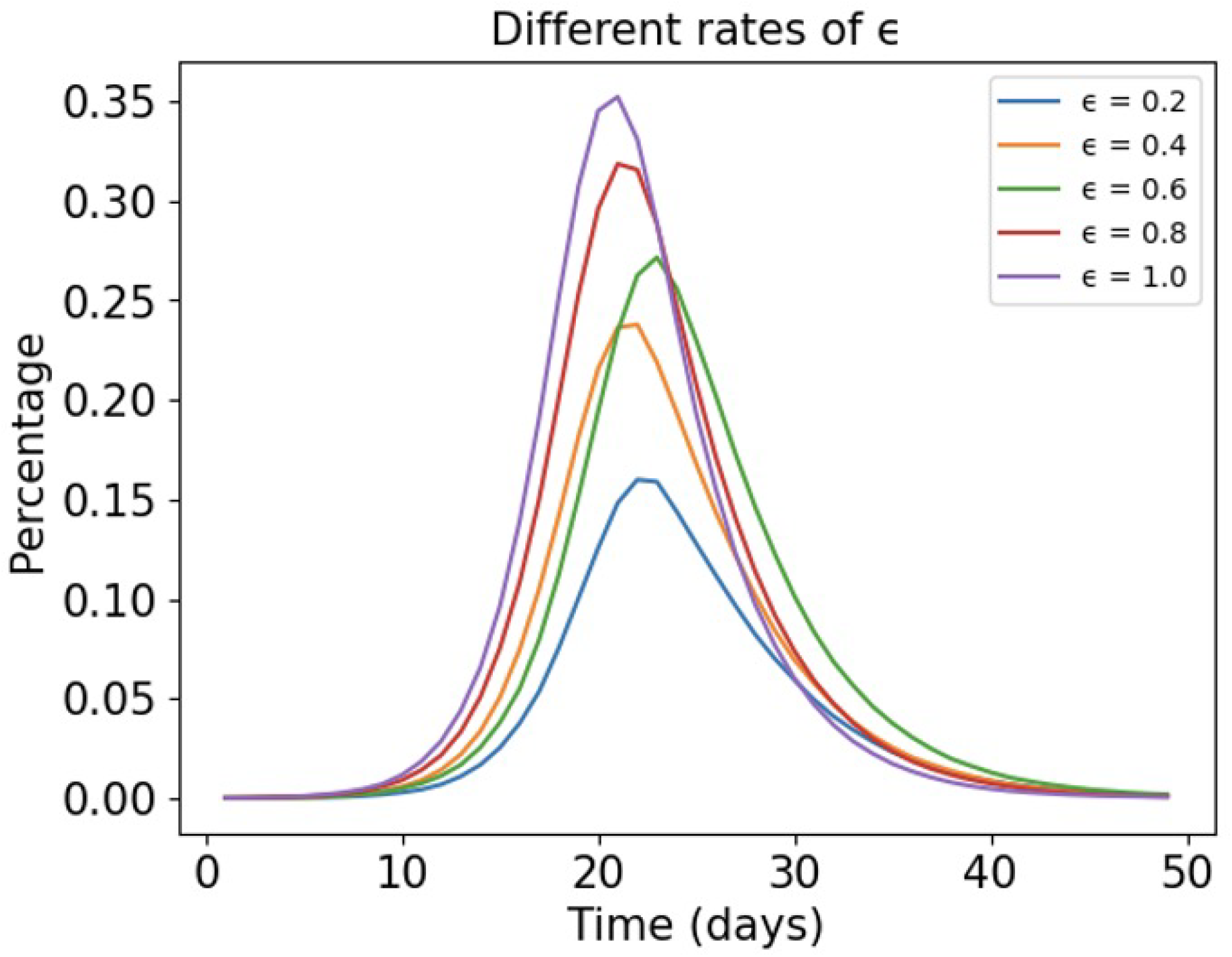
Figure showing the percentage of infected cases in the homeless population over time (in days) under different values of *ϵ* . Note that we set *ν* = 0.002 during the simulation.

- Stronger vaccine with smaller values of *ϵ* can not only reduce the peak of infection rate but also flatten the curve and shorten the outbreak period.
- The case when *ϵ* = 1 means there is no impact on susceptibility from the vaccine, but there may still be benefits from reducing severity. And it is clear that giving homeless vaccine at the beginning of outbreak will greatly mitigate the outbreak in homeless hostels even though the vaccine is less effective, such as *ϵ* = 0.8 .
- From the perspective of controlling outbreak in homeless hostels, it is optimal to give the homeless population the most effective vaccine, which can be important to homeless population as they overall have poorer health conditions and limited medical resources. Reducing infection rate will eventually reduce the cases of hospitalization or mechanical ventilation.

#### B.2.5. Setting with external disease pressure, limited demographic change, no transmission reduction

We assume that *π* is the prevalence of disease in an external community and investigate the impact of this force of infection on a setting. We assume prevalence in community to be approximately constant on the timescale of outbreak in setting (this timescale is defined to be *T*) and *λ* is the contact rate between the setting and the community. Then the key equations are

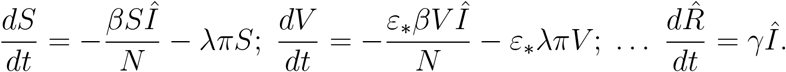

for initial conditions given in Section B.2. As such

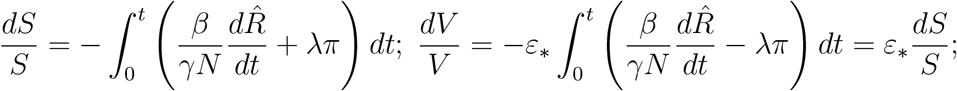

so we may solve 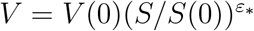 and for some time *T* we have

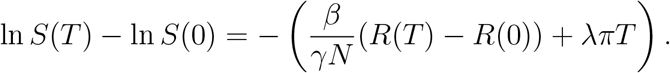

Then if *R*(0) = 0 (no previous natural cases and all immunity is due to vaccination), *S*(0) = (1 − *ω*)*N* − *I*_0_ for *V* (0) = *ωN* (meaning the transient disease states are all zero so introduction is via the external disease pressure), we define *ρ* = *β/γ, R*(*T*) = *WN* and *S*(*T*) = *ZN* then

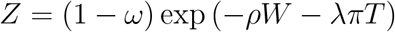

and 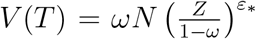. At time *T* we assume that an outbreak has ‘passed’ and so the transient disease states are small relative to *ZN* and *WN* so that the equation of state acts as a further constraint, namely

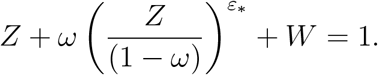

Combining the form for *Z* and equation of state gives

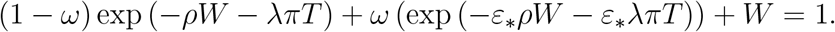

So without vaccine (*ω* = 0) then *Z*_0_ = 1 − *W*_0_ and we have

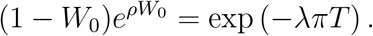

(if *ε*_*\**_ = 1 we return to no impact from vaccine on susceptibility.) So for a desired observed attack ratio post vaccination *W*_*V*_ we have the following formula for effective vaccine coverage Ω

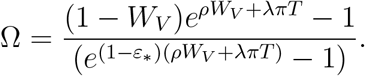

To illustrate the toy model we assume *ε*_*\**_ = 0, *ρ* = 2 ln 2 and *λπT* = 0.5 and aim for *W*_*V*_ = 0.1 then

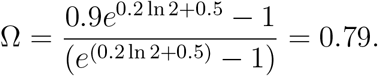

#### B.2.6. How vaccination coverage in community population influences the outbreak in homeless hostels

In the previous scenario, we discussed the impact of vaccination coverage in household population on the outbreak in homeless hostels when the vaccine can reduce the transmission. Here, we change the values of *ν* between 0.0, 0.002, 0.01,0.02, and 0.2. Upon the data from US CDC, recent studies show that flu vaccination reduces the risk of flu illness by between 40% and 60% among the overall population (cf. Centers for Disease Control and Prevention (2023*b*)). Hence, we assume that *ϵ* = 0.5 and *ϕ* = 1 . In Figure (9), we can see that:

**Figure 9:**
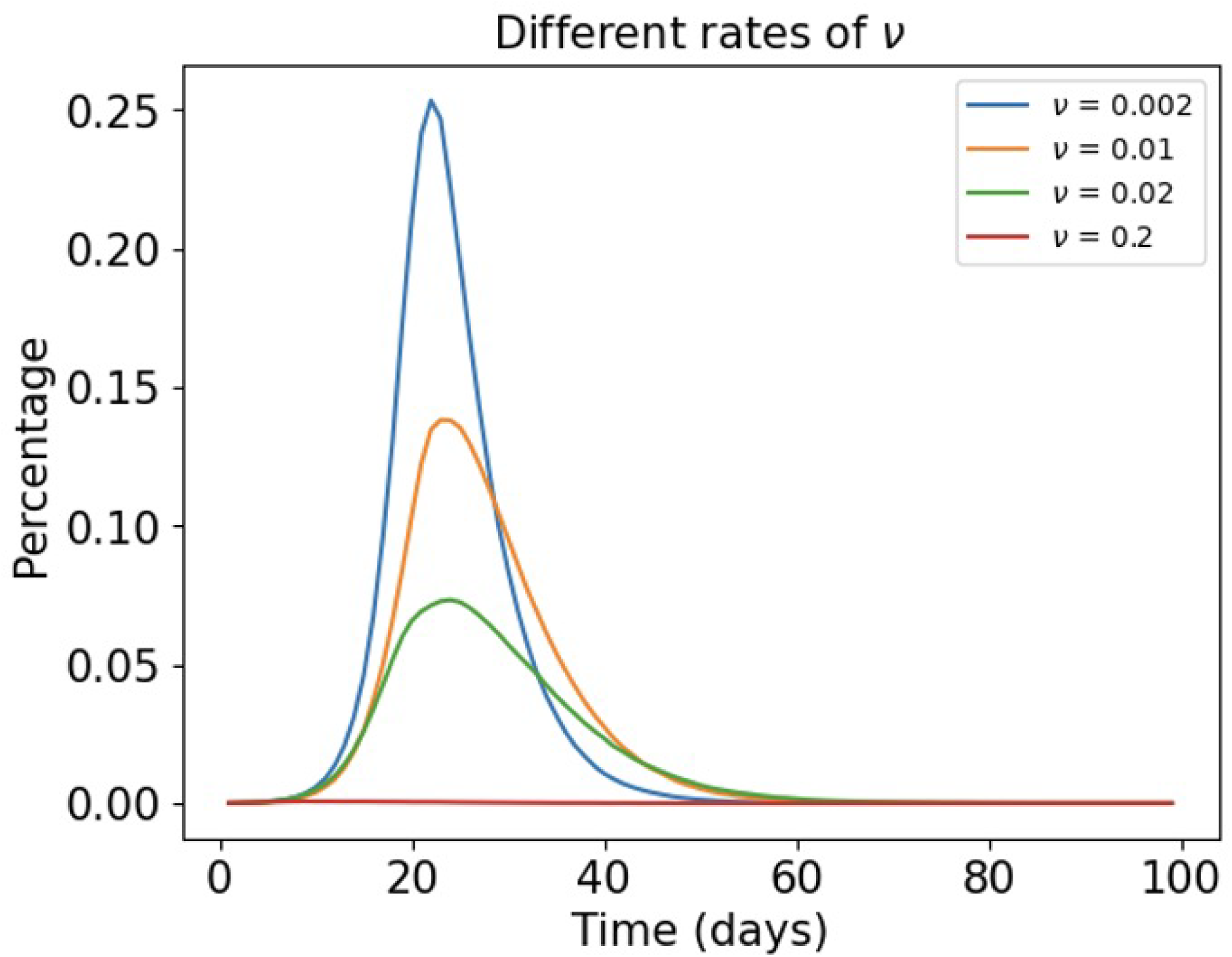
Figure showing the percentage of infected cases in the homeless population over time (in days) under different values of *ν* . Note that we set *ϵ* = 0.5 and *ϕ* = 1 during the simulation.

- Increasing vaccination coverage in household population will reduce the infection rate in homeless hostel and delay the time of peak. The reason is large vaccination coverage in household population will greatly reduce external infection force.
- However, increasing vaccination coverage in homeless hostels can be economically costly, which will be further discussed in the cost and benefit section.

## C. Supplementary Material: Cost of Outbreaks

The outbreak costs *C*_*U*_ and *C*_*V*_ are estimated by adding three separate factors, the personal impact on individual (dominated by the quality adjusted life year), the health care cost (including the primary and secondary care provision) and other societal costs (including sick leave pay, wellbeing gain from not having other disease control, etc.) In the following part, we will fully explain each factor.

A QALY, £Q, is a conversion of the quality of life into terms of monetary value (in this case GB £). Influenza can be asymptomatic in *P*_*A*_ of cases, those that develop symptoms have *T*_*M*_ days of illness so each case costs on average (1 − *P*_*A*_)*T*_*M*_ *Q/*365 .

Furthermore, a proportion *P*_*H*_ of cases will be hospitalised for *T*_*H*_ days (additional to the symptomatic illness) so each hospitalisation costs *T*_*H*_ *Q/*365 additional QALY’s. Or each case can cost *P*_*H*_ *T*_*H*_ *Q/*365 on average.

The fatality ratio of *δ* is assumed double for homeless population compared to the general community due to additional frailty and complexity in their lives. A baseline case fatality ratio for homeless is taken to be 0.1%. Assume that influenza mortality is *L* years earlier than might be expected without illness then each death costs an additional *QL* .

We are assuming here that the quality of life lost from relatively mild illness is equivalent to that lost from hospitalisation or death. Further research could be conducted on homeless population to assess value of quality of life, say through an EQ-5D or similar in general and with respiratory infectious diseases. Note that we do not consider the discounting for the benefit into the future. If we do apply a discounting rate per year, denoted *ε*, then the QALY Q cost of premature death would be *Q* Σ_*i*=0,1,…,(*L−*1)_(1 − *ε*)^*i*^ = *Q*(1 − (1 − *ε*)^*L*^)*/ε* could be applied as a scaling factor. For instance, if *ε* = 0.015 and *L* = 10, then QALY loss would be 9.35Q rather than 10Q under no discounting.

A primary case consultation cost *C*_*P*_ but only *p*_*P*_ symptomatic cases seek primary healthcare. A hospitalisation cost is *C*_*s*_ per day. No cost of Mortuary storage is assumed.

We could include societal costs such as the wellbeing benefit gained by having hostels and shelters open but this will be hard to evaluate with current evidence. As such the benefits of reducing case numbers may be under estimated.

Here we sum these costs to develop a mean per case *µ*_*C*_ (using values in Table 2). The cost of a vaccinated case would be *C*_*V*_ = *µ*_*C*_(1 − *ωθ*)*X*_*V*_ where *X*_*V*_ is the number of cases arising following vaccination (and *θ* is the reduction in severity of those cases for vaccine coverage *ω*) and similarly for unvaccinated outbreak *C*_*U*_ = *µ*_*C*_*X*_*U*_ . Recalling the previous cost inequality, we see that *X*_*U*_ − (1 − *ωθ*)*X*_*V*_ *> C*_*D*_*/µ*_*C*_ where *µ*_*C*_ = (1 − *p*_*A*_)(*p*_*P*_ *C*_*P*_ + *T*_*M*_ *Q/*365) + *p*_*H*_ *T*_*H*_ (*Q* + *C*_*H*_)*/*365 + *δLQ* .

We define *p*_*A*_ = 0.4 (proportion) to be the probability of case being asymptomatic. Through literature review, we notice that in South Africa 44% of infections were asymptomatic (Cohen et al. (2021)) while in UK modelling for general population 50% value is often cited (Scientific Pandemic Influenza Group on Modelling (2018)) but parameter choice here is because the homeless population have higher prevalence of co-morbidities. Duration of mild illness is given by *T*_*M*_ = 3.8 days, which is the infectious period used in Hill et al. (2019) (also cf. McCarthy et al. (2020) and Cauchemez et al. (2004)). It should be noted that the WHO suggests that mild symptoms last up to a week with cough lasting longer World Health Organization (2023), so larger values could be considered.

In homeless populations, when a homeless individual gets infected, they may not actively seek primary healthcare. In the work of Fleming et al. (2000), the RCGP suggest a lower 1 in 10 cases report in the UK but care is needed moving from cases to infections and it maybe lower for population with complex lifestyles. The work of Elwell-Sutton et al. (2017)) shows a lower GP registration rates in risk group. Also, in a study from South Africa 66 infections were medically attended whilst 268 displayed symptoms Cohen et al. (2021). Hence, we define the probability of seeking Primary Healthcare is: *p*_*P*_ = 0.2 . Furthermore, for each individual who needs primary care, the cost of primary care *C*_*P*_, is £56 per person (cf. The King’s Fund (2023)).

For an infected individual, we assume that the probability of hospitalisation is *p*_*H*_ = 0.04 (CDC estimate seasonal flu in general population sees hospitalisation in 1% of cases Centers for Disease Control and Prevention (2023*a*) inflated here given co-morbidities in homeless population). A data linkage study Nilsson et al. (2022) in Denmark between February 2020 and October 2021 found higher risk of Covid-19 hospitalisation (IRR 4·36, 95% CI, 3·09-6·14), intensive care (IRR 3·12, 95% CI 1·29-7·52), and death (MRR 8·17, 95% CI, 3·66-18·25) following a positive SARS-Cov-2 PCR test in people experiencing homelessness compared to the general population so the chosen parameter is likely a lower bound. A study in New York had up to 29 fold increased risk of hospitalisation Miyawaki et al. (2020). We further assume that hospitalised individual will spend around 8 days, i.e. *T*_*H*_ = 8, in the hospital. In fact, the duration of hospitalisation varies by season Moss et al. (2020) so could be longer. Examining NHS Hospital Episode Statistics In financial years 2007-08 to 2010-11, admissions for people experiencing homelessness aged 16 to 64 were on average approximately three times the length of a comparison group (McCormick 2016). Average length of stay in under 65s in the general population with influenza was approximately 5 days (Moss 2020) so setting TH at 8, less than double, could be a conservative estimate.

Each individual under secondary (hospital) care will cost *C*_*H*_ = 350 per day, which is the average general population estimate for 2018 season, though there is evidence of variation by season Moss et al. (2020) and could be higher given co-morbidity/complexity in homeless population). It is worth noting the New York analysis showed that the likelihood of mechanical ventilation was 1.6 fold higher in homeless admissions for flu than in non homeless admitted for flu which likely carries additional cost per day Miyawaki et al. (2020).

The case fatality ratio is taken as *δ* = 0.001 . CDC estimate seasonal flu in general population caused about 5000 deaths in 10 million cases Centers for Disease Control and Prevention (2023*a*) doubled here given co-morbidities in homeless population). A study from Canada suggests this increase could be up to 13 times Hwang (2000). Life years lost by premature death is harder to evaluate. Our baseline value *L* = 10 (years), average age in homeless population is given as mid-forties so *L* could be value could be up to 40 Crisis (2023). Although it is often said that people experiencing homelessness have an average age of death in their 40s or 50s Romaszko et al. (2017) this should not be taken as an average life expectancy – this is because most of this data is based on age of death of people dying whilst experiencing homeless. Because few people remain homeless beyond the age of 60 these statistics do not take account of the many people who move out of homelessness and live for longer. The statistics are also skewed by drug related deaths in the 20-40 age range. Thus a 10 year deficit in life expectancy is likely a lower bound.

The scaling factor on QALY is then ((1 − *p*_*A*_)*T*_*M*_ + *p*_*H*_ *T*_*H*_)*/*365 + *δL* = 2.6*/*365 + 0.01 = 0.0171 compared with Non-QALY costs (1 − *p*_*A*_)*p*_*P*_ *C*_*P*_ + *p*_*H*_ *T*_*H*_ *C*_*H*_ = 118 . We use a QALY based on societal persepctive *Q* = 20000 (£, National Institute for Health and Care Excellence (2015)). Since 2015 there has been 39% inflation whilst there maybe a greater willingness to pay for health gains in the poorest part of the population Robson et al. (2017) and so 30,000 maybe a more viable value. Therefore the total average cost per case is *µ*_*C*_ = (1 − *p*_*A*_)(*p*_*P*_ *C*_*P*_ + *T*_*M*_ *Q/*365) + *p*_*H*_ *T*_*H*_ (*Q/*365 + *C*_*H*_) + *δLQ* = 461.19 (£). Note about £200 of this is the contribution from mortality, £130 from hospital costs and £130 from cases with mild illness.

These baseline values could be refined by including specific ages but given the complexity of mapping biological age to risk age in homeless population we leave this for future research. The most sensitive cost of an outbreak is then the personal cost measured through the QALY and within the contributory factors for this the premature mortality dominates. Given the evidence identified we have tried to be conservative and so the cost of case is likely the lower end of the estimates of actual cost.

From the formula for total average cost per case we can see that hospital care (with direct cost to NHS and QALY to person) contribute less than mild symptom costs and mortality cost (and so greater precision in estimating those values is unlikely to be sensitive). Without the inflation on estimates due to the frailty of the homeless population we would have a cost per case of about £240 which could be an absolute lower limit but further work is needed to understand the impacts of influenza and other respiratory diseases in this population.

Increasing the QALY to 30,000 and allowing 15 years rather than 10 years on the life years lost increases the mortality contribution to £450 per case. Increasing length of stay by factor of 1.6 and adding 100 to cost per day of hospital due to increased use fo high-dependency equipment increases hospital associated costs (personal and care) to £250, and similarly the mild cases would cost £200 so the cost per case on average would become £900 effectively doubling the conservative baseline values.

## D. Supplementary Material: Stochastic model vaccine coverage sensitivity analysis

**Figure 10:**
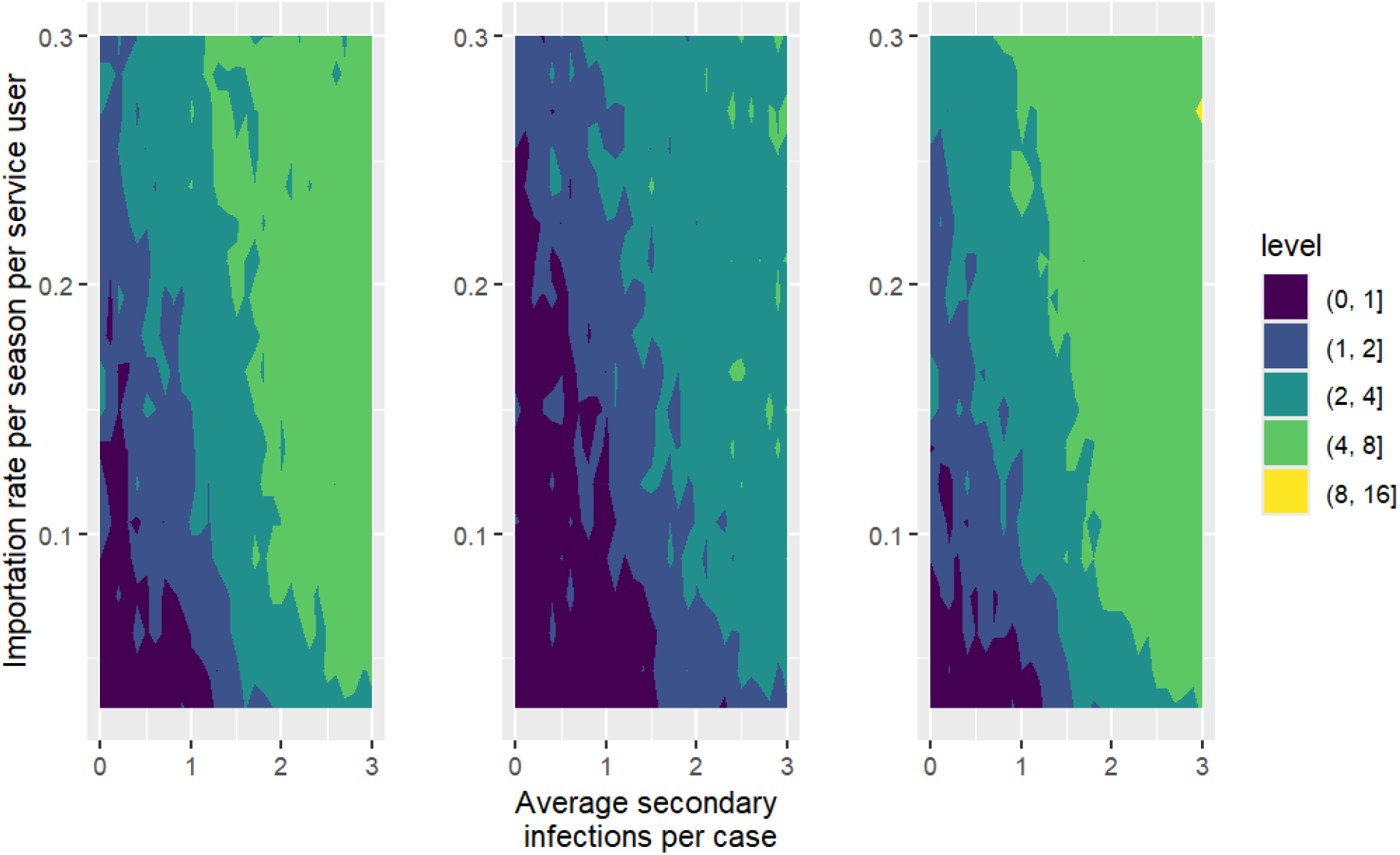
Mean cost effectiveness arising from simulation of outbreaks in 1208 homeless settings assuming vaccine coverage of 50% assuming vaccine is severity reducing only (left panel), susceptibility reducing (middle panel) and both (right panel). The *x* -axis in each panel is the control reproduction number in setting and the *y* -axis the ingress rate of cases from community. The levels marked by contours show the relative mean cost (total cost of cases averted in outbreaks given vaccination divided by total cost of campaign). Those parameter combinations that were not cost effective are marked in darkest colour (levels less than 1 on legend scale).

**Figure 11:**
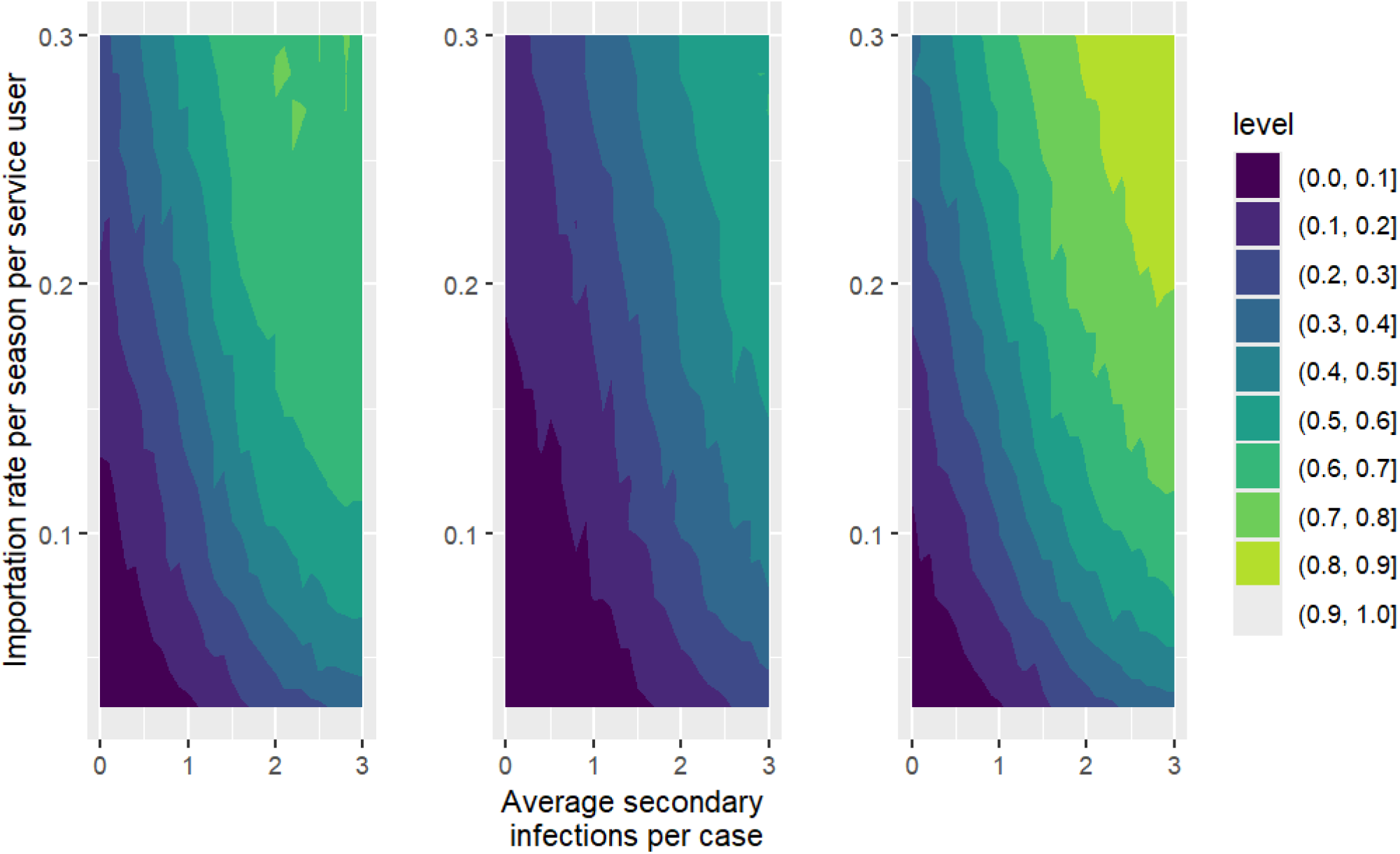
Sample quantile of the simulations that were cost effective for 1208 homeless settings assuming vaccine coverage of 50% assuming vaccine is severity reducing only (left panel), susceptibility reducing (middle panel) and both (right panel). The *x* -axis in each panel is the control reproduction number in setting and the *y* -axis the ingress rate of cases from community, the levels marked by contours show the proportion of simulations that were cost effective

**Figure 12:**
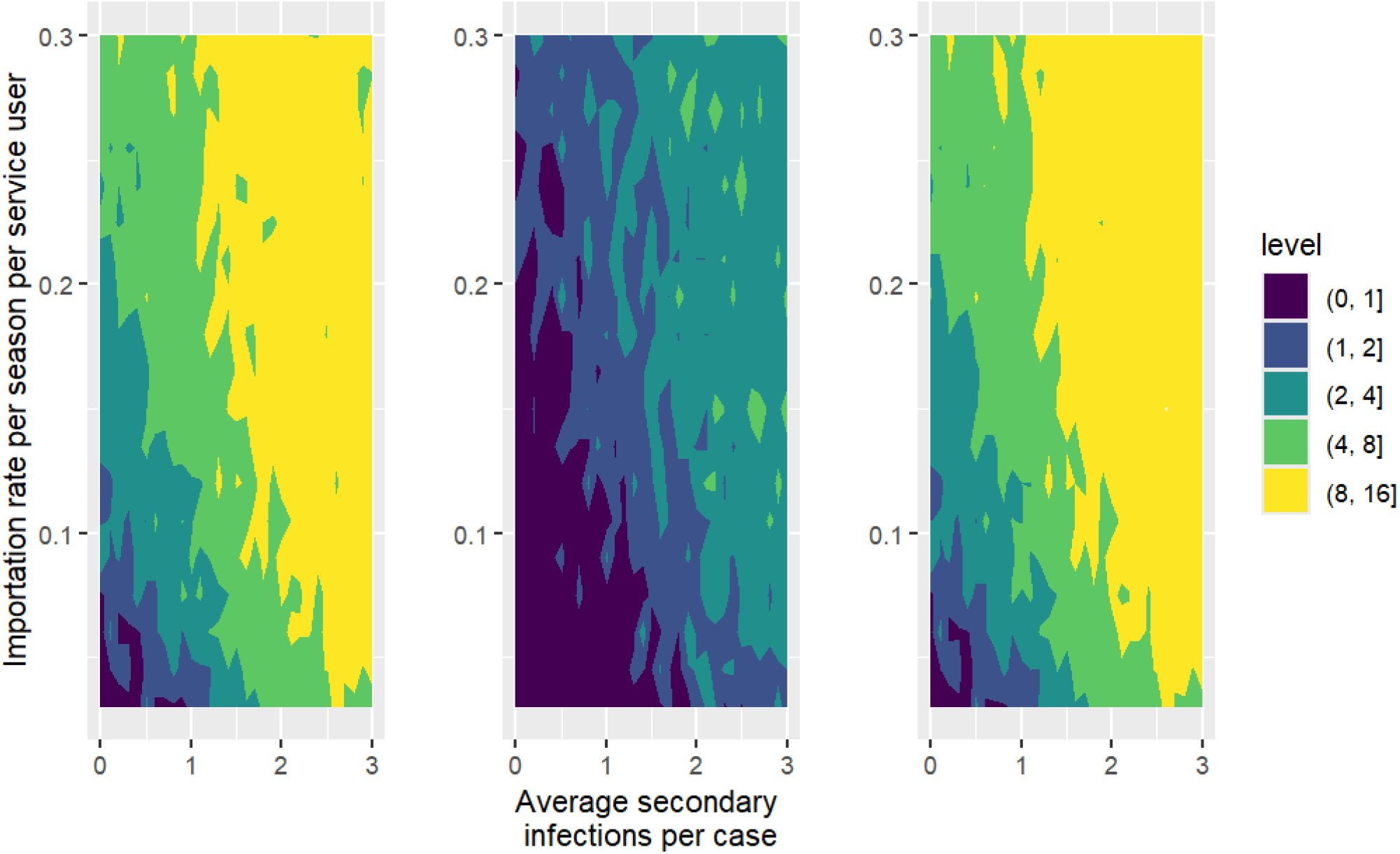
Mean cost effectiveness arising from simulation of outbreaks in 1208 homeless settings assuming vaccine coverage of 25% assuming vaccine is severity reducing only (left panel), susceptibility reducing (middle panel) and both (right panel). The *x* -axis in each panel is the control reproduction number in setting and the *y* -axis the ingress rate of cases from community. The levels marked by contours show the relative mean cost (total cost of cases averted in outbreaks given vaccination divided by total cost of campaign). Those parameter combinations that were not cost effective are marked in darkest colour (levels less than 1 on legend scale).

**Figure 13:**
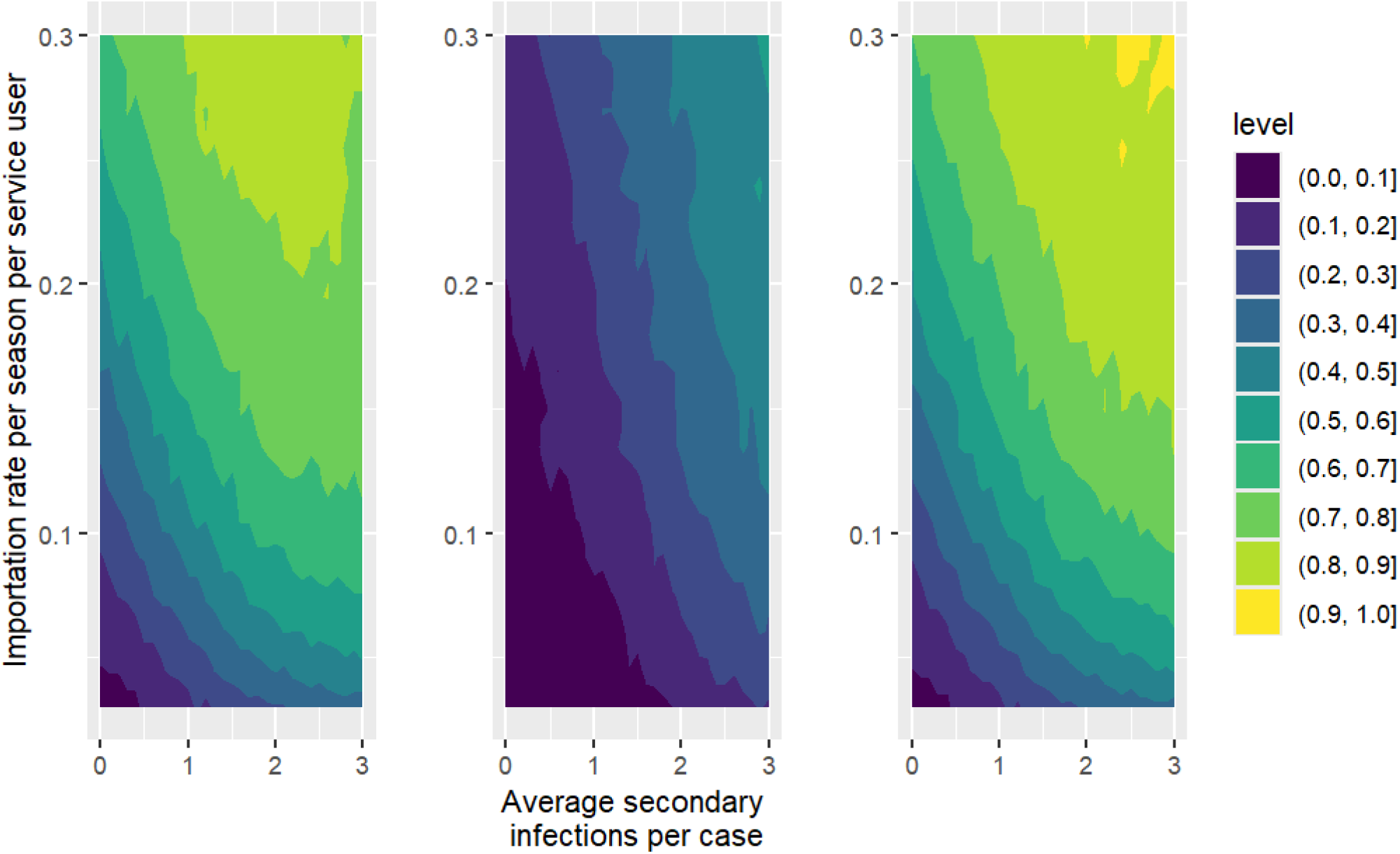
Sample quantile of the simulations that were cost effective for 1208 homeless settings assuming vaccine coverage of 25% assuming vaccine is severity reducing only (left panel), susceptibility reducing (middle panel) and both (right panel). The *x* -axis in each panel is the control reproduction number in setting and the *y* -axis the ingress rate of cases from community, the levels marked by contours show the proportion of simulations that were cost effective

